# Natural Anti-NMDAR1 Autoantibodies Associate with Slowed Decline of Cognitive Functions in Alzheimer’s Diseases

**DOI:** 10.1101/2025.01.16.25320688

**Authors:** Xianjin Zhou

## Abstract

Anti-NMDAR1 autoantibodies can bind NMDA receptors to suppress glutamate excitotoxicity in the brain. Low titers of blood circulating natural anti-NMDAR1 autoantibodies were reported in ∼10% of the general human population. We developed a new method to more accurately quantify these low titers of natural anti-NMDAR1 autoantibodies. After quantifying natural anti-NMDAR1 autoantibodies in the plasma of 324 age– and sex-matched subjects (163 healthy controls; 161 Alzheimer’s disease (AD) patients), I found that AD patients carrying higher levels of natural anti-NMDAR1 autoantibodies have significantly (p value: 0.003) higher scores of Mini-Mental State Examination (MMSE score: 23.5) than AD patients carrying lower levels of natural anti-NMDAR1 autoantibodies (MMSE score: 21.4). No significant differences in MMSE scores were however found between healthy controls with either higher or lower levels of natural anti-NMDAR1 autoantibodies, indicating little harmful effect of the autoantibodies. Consistently, superior cognitive performances were found in AD patients carrying higher levels of natural anti-NMDAR1 autoantibodies in comparison with AD patients carrying lower levels of the autoantibodies. These data suggest that natural anti-NMDAR1 autoantibodies may have neuroprotective effects against cognitive decline in AD patients.

## Introduction

Alzheimer’s disease (AD) is the most common dementia and affects ∼ 5.8 million Americans. AD treatment costed about $305 billion in 2020 and is expected to cost more than $1 trillion as the population ages. Monoclonal antibodies against amyloid-β were recently approved by FDA as disease-modifying therapies for AD with modest and inconsistent effects on slowing down cognitive decline (Budd Haeberlein et al., 2022; van Dyck et al., 2023). Potential life-threatening side effects can however occur (Niazi, 2024). More effective and safer treatments to slow down cognitive decline in AD patients are needed.

Different hypotheses for AD development were proposed as reviewed (Zhang et al., 2024). Glutamate excitotoxicity is one of the potential mechanisms involved in cognitive decline during AD development (Hynd et al., 2004). In fact, glutamate excitotoxicity is a common mechanism for neuronal injury in both neurological diseases (Lai et al., 2014) (e.g. stroke, epilepsy) and neurodegenerative diseases (Binvignat and Olloquequi, 2020) (e.g. AD, PD, HD, ALS), and also contributes to the pathogenesis of psychiatric disorders such as schizophrenia (Kraguljac et al., 2013; Zhou et al., 2005). In Alzheimer’s disease, amyloid-β induces glutamate release from both neuronal and glial cells to cause a slow buildup of extracellular glutamate (Li et al., 2011; Talantova et al., 2013) that activates extrasynaptic NMDARs to further increase amyloid-β production (Bordji et al., 2010) and tau over-expression (Sun et al., 2016), indicating an important role for glutamate excitotoxicity mediated by extrasynaptic NMDARs in exacerbating AD pathogenesis.

Suppression of glutamate excitotoxicity was demonstrated with anti-NMDAR1 autoantibodies in animal models of stroke and epilepsy (During et al., 2000). Natural anti-NMDAR1 autoantibodies were reported in the blood of ∼5-10% of the general human population using semi-quantitative methods (Castillo-Gomez et al., 2017; Hammer et al., 2014; Jezequel et al., 2017; Pan et al., 2019). We developed a new method for more accurate quantification of blood natural anti-NMDAR1 autoantibodies (Vaughn et al., 2024). I am wondering whether blood natural anti-NMDAR1 autoantibodies may ameliorate glutamate excitotoxicity after crossing blood-brain barriers during AD development. Using our recently developed new methodology, I quantified the levels of blood natural anti-NMDAR1 autoantibodies in both healthy controls and early-stage AD patients. I found that AD patients carrying higher levels of natural anti-NMDAR1 autoantibodies have significantly better cognitive performances than AD patients carrying lower levels of natural anti-NMDAR1 autoantibodies.

## Materials and Methods

### Healthy Controls and Patients with Alzheimer’s Disease

EDTA plasma samples from Alzheimer’s disease patients (n=161) and healthy controls (n=163) were received from Alzheimer’s Disease Research Center (ADRC) at University of California San Diego (UCSD) with an IRB Protocol 170957. The plasma samples were predominantly from the first visit of the subjects. All subjects are with ages over 60; and most were examined with Mini-Mental State Examination (MMSE), Clinical Dementia Ranking Sum (CDRSUM), and some cognitive tests.

### Quantitative Immunoassay

Both *Gaussia* luciferase (GLUC) and NMDAR1-GLUC fusion proteins were purified as described (Vaughn et al., 2024) as the probes for quantification of plasma natural anti-NMDAR1 autoantibodies. Natural anti-NMDAR1 autoantibodies in human plasma samples were quantified using our recently developed method (Vaughn et al., 2024). A negative control with goat serum was used as background. Three positive controls with different levels of anti-NMDAR1 autoantibodies were used as references for normalization between different 96-well plates (Greiner 96-well Flat Bottom Black Polystyrene plate, Cat. No.: 655097). Each human plasma sample was supplemented with the same amount of goat serum as in the negative control for the immunoassay.

*Gaussia* luciferase substrate (ThermoFisher, cat. 16160; Pierce™ *Gaussia* Luciferase Glow Assay Kit) was used for quantification on Tecan infinite 200Pro. After subtracting the background Relative Light Units (RLU) of the negative control, the remaining RLU will be the levels of natural anti-NMDAR1 autoantibodies in individual plasma samples. A ratio between the RLU of each plasma sample against the average RLU within the same plate will be used as the relative levels for ranking the autoantibodies.

### Statistical Analysis

ANOVA was conducted for all statistical analyses using R programming. Welch’s t-test was used as a post-hoc analysis to compare the means of two groups.

## Results

I quantified natural anti-NMDAR1 autoantibodies in the plasma of 324 subjects including 164 healthy controls and 161 AD patients. There is no age difference between the control and the patient groups (F1(1,320)=2.655, p=0.104) or between the genders (F(1,320)=0.986, p=0.321) (Figure 1A). However, There is an interaction between gender and diagnosis on the levels of natural anti-NMDAR1 autoantibodies (F(1,320)=4.15, p=0.0425) (Figure 1B). *Post hoc* analysis revealed significant higher levels of natural anti-NMDAR1 autoantibodies in the plasmas of male AD patients than male controls (Welch’s *t*-test, t= 2.2992, p=0.02323). There is no APOE effect (F(6,305)=0.57, p=0.7545) on the levels of natural anti-NMDAR1 autoantibodies (Figure 2A). To investigate age effect, I separated all subjects into 4 different groups according to their ages (Figure 2B). No significant age effect was observed (F(1,316)=2.375, p=0.1243) on the levels of natural anti-NMDAR1 autoantibodies.

**Figure 1.**
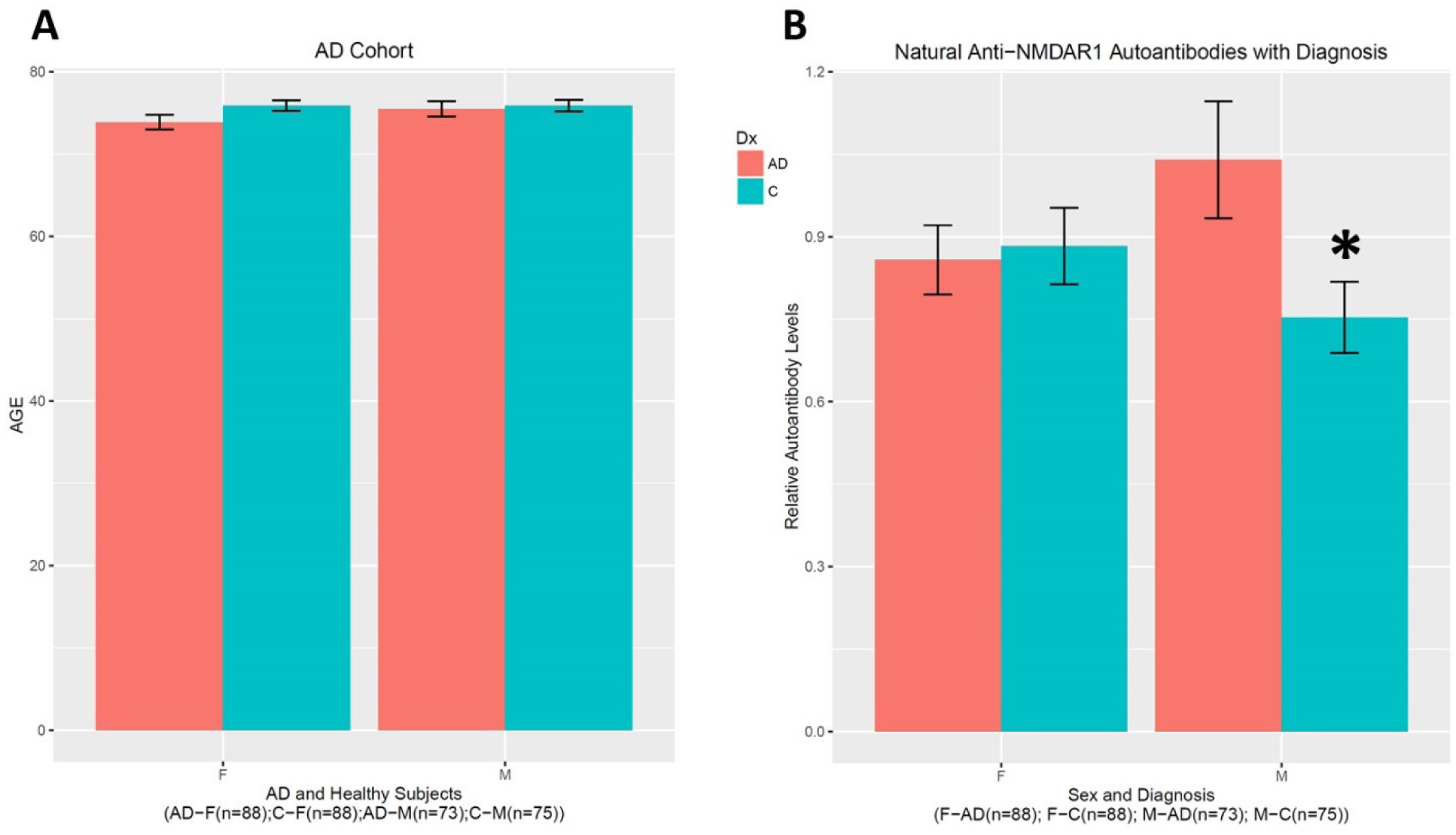
Higher levels of natural anti-NMDAR1 autoantibodies in male AD patients. **(A)** There is no age difference in between male and female subjects or between healthy controls and AD patients. **(B)** An interaction between gender and AD (F(1,320)=4.15, p=0.0425) was found on the levels of natural anti-NMDAR1 autoantibodies. A significant higher level of natural anti-NMDAR1 autoantibodies was observed in the plasmas of male AD patients than in the plasma of male controls (Welch’s *t*-test, t= 2.2992, p=0.02323). Data were presented as Mean+SEM. *p* value: * < 0.05.

**Figure 2.**
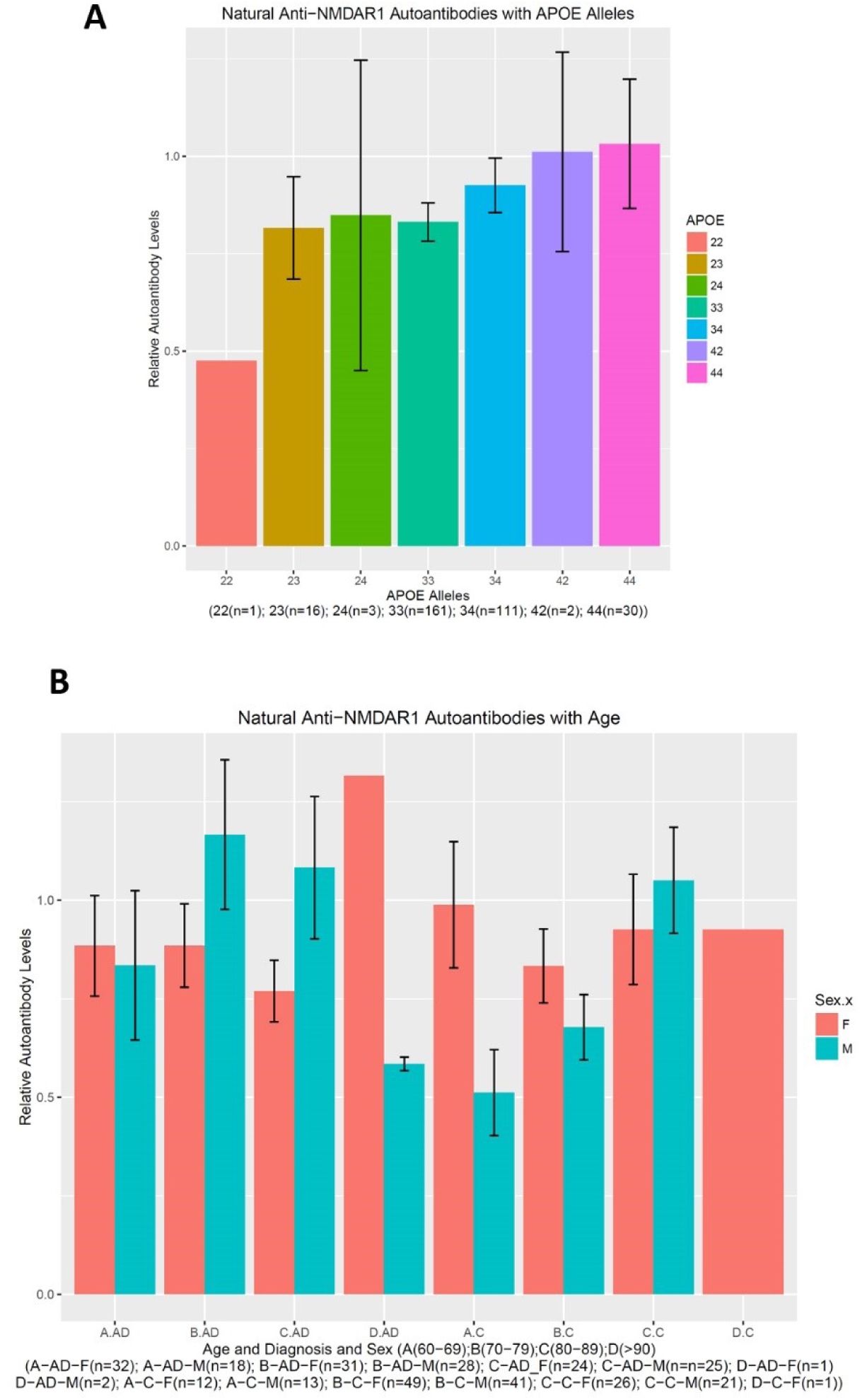
Neither APOE alleles nor age affects the levels of natural anti-NMDAR1 autoantibodies. **(A)** There are many different APOE alleles in the cohort. No effect of APOE alleles were observed (F(6,305)=0.57, p=0.7545) on the levels of natural anti-NMDAR1 autoantibodies. **(B)** All subjects were over 60-year-old and were separated into 4 different age groups (A: 60-69; B: 70-79; C: 80-89; D: >90). No significant age effect was observed (F(1,316)=2.375, p=0.1243) on the levels of natural anti-NMDAR1 autoantibodies between the 4 age groups. Data were presented as Mean+SEM.

To investigate effects of blood natural anti-NMDAR1 autoantibodies on cognitive functions, I ranked subjects according to their autoantibody levels and arbitrary assigned the top quarter of the autoantibody levels as the “High” group, the rest (75%) as the “Low” group. A significant interaction between diagnosis and the autoantibody (F(1,313)=7.698, p=0.00586) was observed on the MMSE scores (Figure 3A). *Post hoc* analysis revealed significantly higher MMSE scores in AD patients carrying higher levels of natural anti-NMDAR1 autoantibodies than in AD patients carrying lower levels of natural anti-NMDAR1 autoantibodies (Welch’s *t*-test, t= 2.9848, p=0.003583). No gender effect (F(1,313)=0.074, p=0.785) or interactions between gender and diagnosis (F(1,313)=1.024, p=0.3122) or between gender and the autoantibody (F(1,313)=1.201, p=0.274) were observed on MMSE scores. There is no autoantibody effect (F(1,304)=0.567, p=0.452) or interaction between the autoantibody and diagnosis (F(1,304)=0.330, p=0.566) on Clinical Dementia Ranking Sum (CDRSUM)(Figure 3B).

**Figure 3.**
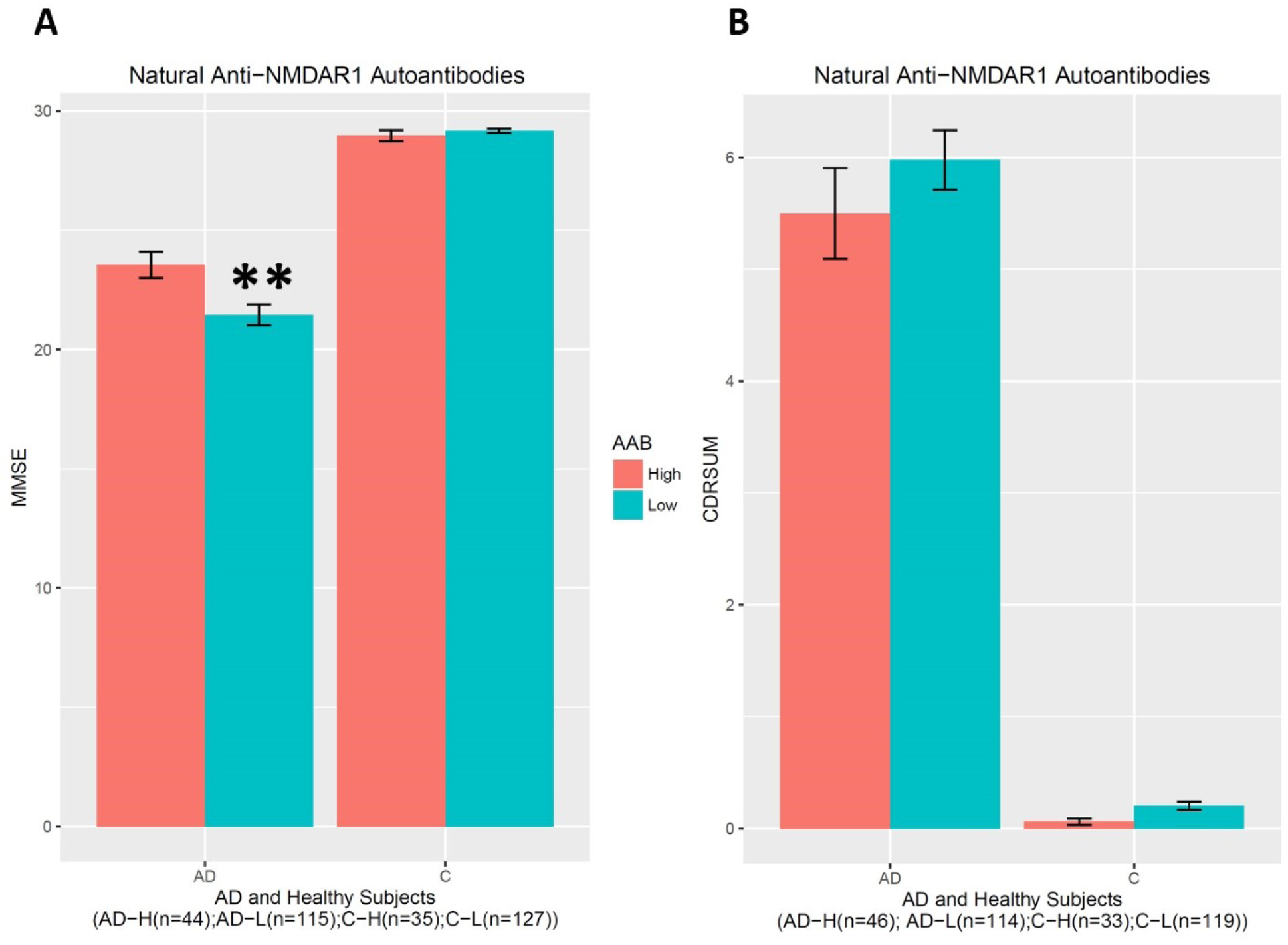
Higher levels of natural anti-NMDAR1 autoantibodies are associated with slowed decline of MMSE scores in AD patients. **(A)** No gender (F(1,313)=0.074, p=0.785) or autoantibody (F(1,313)=1.201, p=0.274) effect or interactions between gender and AD (F(1,313)=1.024, p=0.3122) on MMSE scores. A significant interaction between diagnosis and the autoantibody (F(1,313)=7.698, p=0.00586) was observed on the MMSE scores. *Post hoc* analysis revealed significantly higher MMSE scores in AD patients carrying higher levels of natural anti-NMDAR1 autoantibodies than AD patients carrying lower levels of natural anti-NMDAR1 autoantibodies (Welch’s *t*-test, t= 2.9848, p=0.003583). **(B)** There is no significant effect on CDRSUM by natural anti-NMDAR1 autoantibodies. Data were presented as Mean+SEM. *p* value: ** < 0.01.

Many neuropsychologic tests were performed for this cohort, but many subjects have missing data. A subset of these tests was therefore analyzed. There is a significant gender effect on Verbal Fluency Correct Response (F(1,311)=5.044, p=0.025) (Figure 4A) and Verbal Fluency Total Words (F(1,311)=4.924, p=0.027) (Figure 4B). A significant interaction between diagnosis and the autoantibody was observed in either Verbal Fluency Correct Response (F(1,311)=7.528, p=0.00643) or Verbal Fluency Total Words (F(1,311)=7.543, p=0.00638). *Post hoc* analysis revealed significantly better performances in AD patients carrying higher levels of natural anti-NMDAR1 autoantibodies than in AD patients carrying lower levels of natural anti-NMDAR1 autoantibodies in either Verbal Fluency Correct Response (In female AD patients, Welch’s *t*-test, t= 2.1227, p=0.04; in male AD patients, Welch’s *t*-test, t= 2.0938, p=0.042) or Verbal Fluency Total Words (In female AD patients, Welch’s *t*-test, t=1.8743, p=0.069; in male AD patients, Welch’s *t*-test, t= 2.253, p=0.029). For short-term memory and learning, significant interactions between diagnosis and the autoantibody were observed in both WAIS-R Digital Symbol Test (F(1,284)=6.614, p=0.01) (Figure 5A) and Scaled WAIS-R Digital Symbol Test (F(1,283)=6.214, p=0.0132) (Figure 5B). *Post hoc* analysis revealed significantly better performances in AD patients carrying higher levels of natural anti-NMDAR1 autoantibodies than in AD patients carrying lower levels of natural anti-NMDAR1 autoantibodies in either WAIS-R Digital Symbol Test (Welch’s *t*-test, t= 2.4684, p=0.016) or Scaled WAIS-R Digital Symbol Test (Welch’s *t*-test, t= 2.4778, p=0.0157).

**Figure 4.**
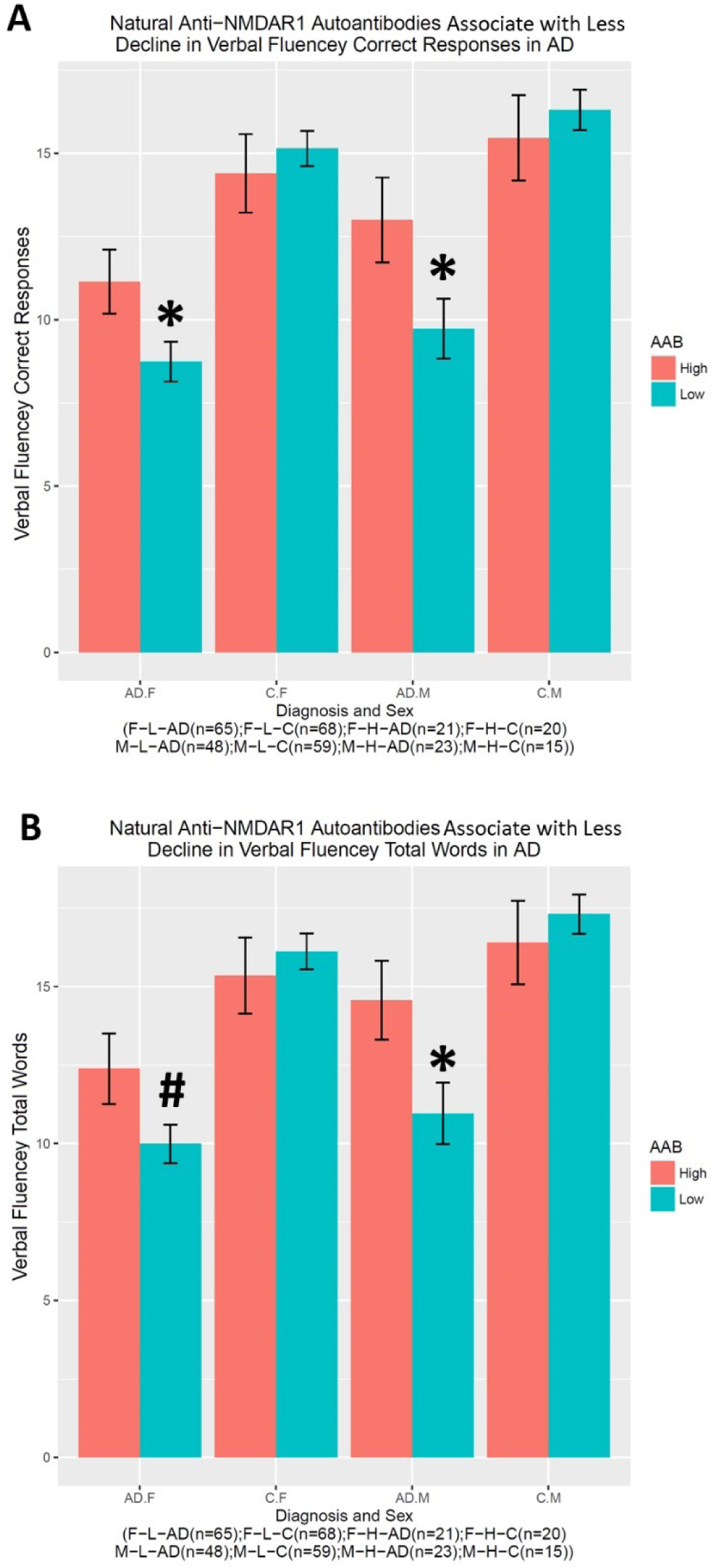
Higher levels of natural anti-NMDAR1 autoantibodies are associated with slowed decline of verbal fluency in AD patients. There is a significant gender effect between male and female subjects on either verbal fluency correct response (F(1,311)=5.044, p=0.025) **(A)** or verbal fluency total words (F(1,311)=4.924, p=0.027) **(B)**. Significant interactions between diagnosis and autoantibody were observed in both Verbal Fluency Correct Response (F(1,311)=7.528, p=0.00643) **(A)** and Verbal Fluency Total Words (F(1,311)=7.543, p=0.00638) **(B)**. *Post hoc* analyses revealed significantly better performances in both verbal fluency correct responses and total words in AD patients carrying higher levels of natural anti-NMDAR1 autoantibodies than AD patients carrying lower levels of natural anti-NMDAR1 autoantibodies. Data were presented as Mean+SEM. *p* value: ^#^ < 0.1; * < 0.05.

**Figure 5.**
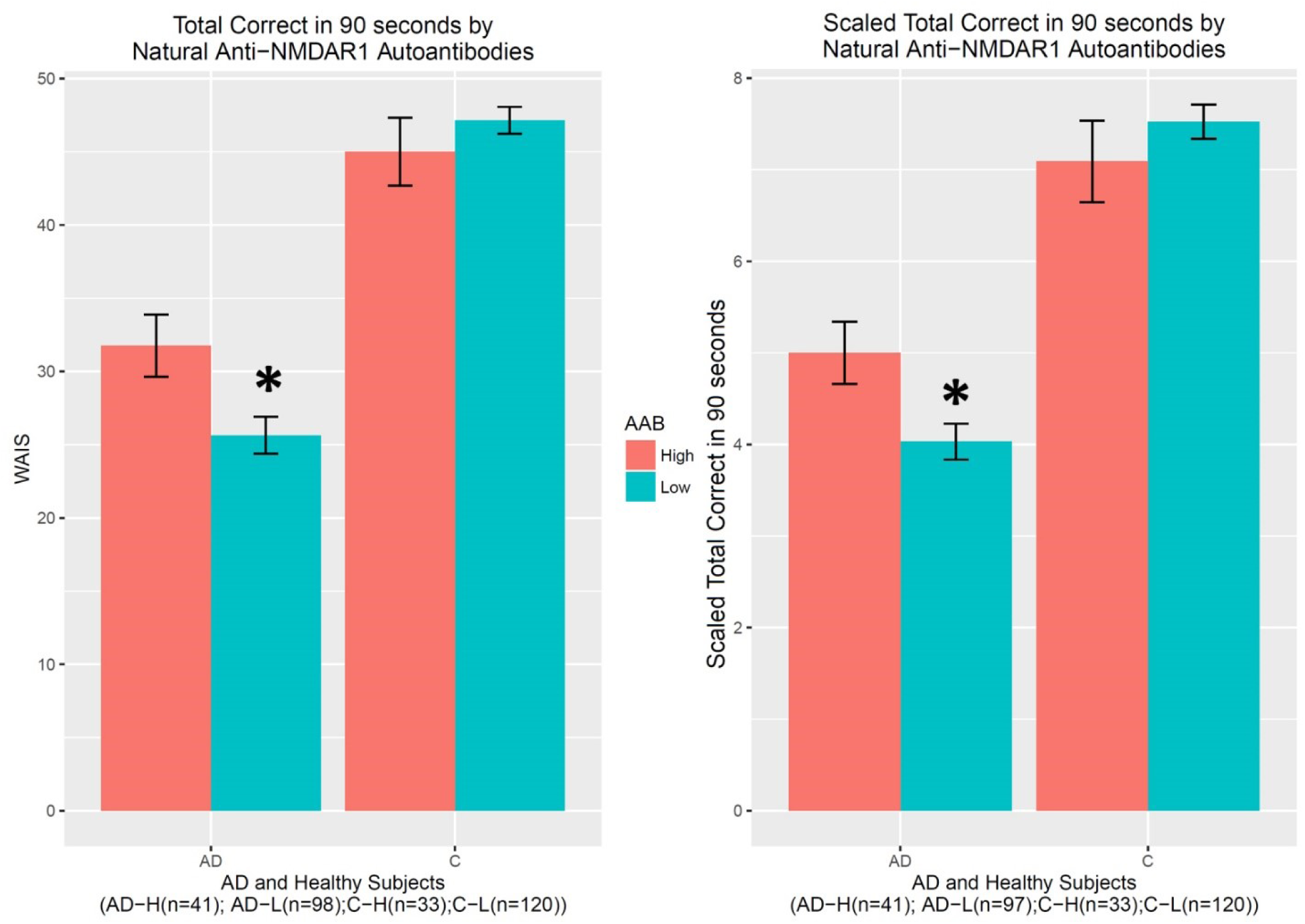
Higher levels of natural anti-NMDAR1 autoantibodies are associated with slowed decline in short-term memory and learning in AD patients. There is no gender effect on WAIS-R Digital Symbol Test for short-term memory and learning. A significant interaction between diagnosis and the autoantibody was observed in either WAIS-R Digital Symbol Test (F(1,284)=6.614, p=0.01) **(A)** or Scaled WAIS-R Digital Symbol Test (F(1,283)=6.214, p=0.0132) **(B)**. *Post hoc* analyses revealed significantly better performances in either WAIS-R Digital Symbol Test (Welch’s t-test, t= 2.4684, p=0.016) or Scaled WAIS-R Digital Symbol Test (Welch’s t-test, t= 2.4778, p=0.0157) in AD patients carrying higher levels of natural anti-NMDAR1 autoantibodies.

To investigate how much non-specific binding from plasma overall general antibodies may contribute to RLU readings of natural anti-NMDAR1 autoantibodies, I prepared a new GLUC probe without NMDAR1 fusion to assess the levels of non-specific bindings in randomly selected plasma samples carrying different levels of natural anti-NMDAR1 autoantibodies. Since our method can conduct cross-species quantification of anti-NMDAR1 autoantibodies (Vaughn et al., 2024), I used control mice and immunized mice carrying anti-NMDAR1 autoantibodies (Yue et al., 2021) as the controls for quantification (Figure 6). With the same input of GLUC or NMDAR1-GLUC luciferase activities, there is a very low amount of precipitated GLUC activities across all mouse serum and human plasma samples regardless of the levels of anti-NMDAR1 autoantibodies measured by the NMDAR1-GLUC probe. This data suggested that anti-NMDAR1 autoantibodies were specifically measured for individual plasma samples rather than non-specific bindings from plasma overall general antibodies.

**Figure 6.**
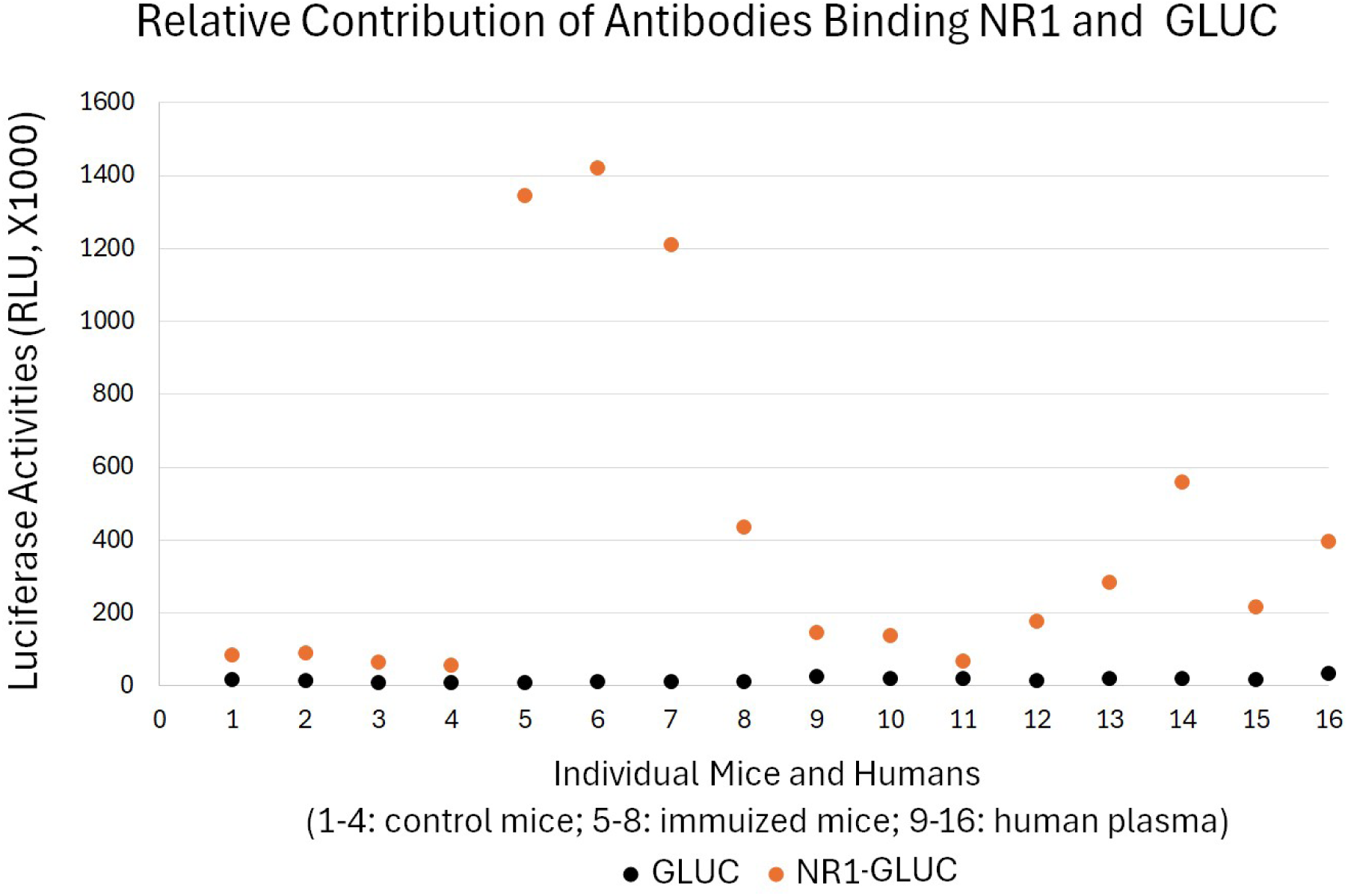
Little contribution from non-specific background binding to high RLU readings of anti-NMDAR1 autoantibodies. Specificity of the quantification of natural anti-NMDAR1 autoantibodies were validated in both immunized mice and human subjects using both GLUC and NR1-GLUC probes. The same amount of input GLUC and NR1-GLUC luciferase activities were used for quantification using exactly the same protein AGL mixture to assess both non-specific background binding and specific anti-NMDAR1 antibody binding across mouse and human. Control wildtype mice (1-4); immunized mice carrying anti-NMDAR1 autoantibodies (5-8). Randomly selected human subjects carrying different levels of natural anti-NMDAR1 autoantibodies (9-16).

## Discussion

High titers of anti-NMDAR1 IgG autoantibodies in brain can cause anti-NMDAR1 encephalitis that exhibits psychosis, impaired memory, and many other psychiatric symptoms in addition to neurological symptoms (Dalmau, 2016). We previously found that low titers of blood circulating anti-NMDAR1 IgG autoantibodies impair spatial working memory in mice with intact blood-brain barriers (Yue et al., 2021). However, our cross-species quantification revealed that the levels of natural anti-NMDAR1 autoantibodies in human plasma are lower than the levels of anti-NMDAR1 autoantibodies generated in immunized mice (Yue et al., 2021). Importantly, cognitive performances of healthy human controls carrying higher levels of natural anti-NMDAR1 autoantibodies are comparable to the controls carrying lower levels of natural anti-NMDAR1 autoantibodies, suggesting little harmful effect of plasma natural anti-NMDAR1 autoantibodies.

More importantly, higher levels of plasma natural anti-NMDAR1 autoantibodies are associated with slower cognitive decline in AD patients as shown in MMSE and other cognitive tests, suggesting that natural anti-NMDAR1 autoantibodies may be neuroprotective. Given that blood anti-NMDAR1 autoantibodies suppress glutamate excitotoxicity in animal models of stroke and epilepsy (During et al., 2000), it is plausible that plasma natural anti-NMDAR1 autoantibodies may ameliorate glutamate excitotoxicity in brain during AD development. About 0.1% of blood circulating antibodies nonspecifically cross blood-brain barriers into brain in healthy rodents and humans regardless of antibody specificities or isotypes (Banks, 2010; Banks et al., 2007; Wang et al., 2018). Blood-brain barriers are compromised in AD patients (Sweeney et al., 2018), which may facilitate crossover of plasma anti-NMDAR1 autoantibodies into brain. Since natural anti-NMDAR1 autoantibodies persist for months and years, basal crossover of natural anti-NMDAR1 autoantibodies into brain parenchyma may be sufficient to provide neuroprotective effects via suppressing glutamate excitotoxicity.

Glutamate excitotoxicity is mainly caused by excessive glutamate to activate extrasynaptic NMDA receptors (NMDARs) localized outside of the synapse, whereas activation of synaptic NMDARs promotes neuronal survival and mediates synaptic transmission for cognitive functions (Karpova et al., 2013; Wu and Tymianski, 2018).

Blood natural antibodies are mainly IgM and IgA isotypes. I proposed that that anti-NMDAR1 IgM autoantibodies may specifically inhibit extrasynaptic NMDARs to suppress glutamate excitotoxicity, but spare synaptic NMDARs, because the IgM autoantibodies are physically too large to enter the synaptic cleft (Zhou, 2023).

Consistent with my hypothesis, memantine that is an NMDAR antagonist preferentially (but not specifically) inhibiting extrasynaptic NMDARs (Xia et al., 2010) has been approved by FDA as a treatment for Alzheimer’s disease. In contrast to IgM, IgG anti-NMDAR1 autoantibodies can access synaptic NMDARs due to their small sizes and thereby may inhibit synaptic NMDA neurotransmission to compromise cognitive functions. Quantification of IgM isotype anti-NMDAR1 autoantibodies in this cohort of AD patients and healthy controls will therefore be important to validate the hypothesis. Validation of the hypothesis may open a new avenue for development of the IgM therapeutics for AD patients.

Females have higher incidence rates of developing AD than males at the same old ages (Beam et al., 2018) and progress at faster rates than males after AD diagnosis (Lin et al., 2015). Interestingly, male AD patients have significant higher levels of natural anti-NMDAR1 autoantibodies than healthy male controls, but not than female AD patients yet (*p*=0.14). A larger cohort of plasma samples is needed to confirm if male AD patients carry higher levels of natural anti-NMDAR1 autoantibodies than female AD patients to provide more neuroprotection, which may contribute to gender differences in AD development.

## Data Availability

All data produced in the present study are available upon reasonable request to the authors

## Acknowledgements

All plasma samples were obtained from Alzheimer’s Disease Research Center (ADRC) at University of California San Diego (UCSD), which was supported by the National Institute on Aging of the National Institutes of Health under award number: P30 AG062429. I would like to thank Douglas Galasko, Timothy Gahagan, and Olivia Ott at ADRC for their help in selection of plasma samples and organization of neuropsychological data.

